# Usability testing with a prototype user interface of an Artificial Intelligence driven air-Safety Tool (AISaT)

**DOI:** 10.64898/2026.06.15.26355448

**Authors:** Sigrún Eyrúnardóttir Clark, Ryo Torii, Yiqing Li, Sanjoli Mathur, Yolanda Barrado-Martín, Fiona Stevenson, Zarnie Khadjesari, Laurence B. Lovat, Cecilia Vindrola-Padros

## Abstract

Involving end-users in the development of an AI tool is an important facilitator to its implementation. Usability testing was therefore conducted with a prototype user interface of an Artificial Intelligence driven air-Safety Tool (AISaT) to capture the perspectives and user experiences of AISaT from 10 staff members across two hospitals working within estates, infection prevention and control, and clinical areas, to inform the development of next iterations of AISaT. The perspectives shared could be grouped under improvements to the understand-ability; content; navigation; visibility; usability; workflow; ownership; and frequency of use of the tool. There were key areas that can and will be easily improved within AISaT, however there were areas that required a deeper level of critical reflection, such as incorporating data on more existing variables in a room (i.e., existing ventilation) and determining who is infected, and the level of breathing.

## Introduction

The implementation of artificial intelligence (AI) within healthcare settings is growing globally [1], especially within the UK context where there is strong support from the Government to make ‘the NHS the most artificial-intelligence-enabled care system in the world’ [2]. AI encompasses computational systems that can use algorithms to perform pattern recognition and problem solving [3,4]. An early-stage tool developed by a research team at University College London, has incorporated AI and computational fluid dynamics (CFD). CFD combines the disciplines of fluid mechanics, mathematics and computer science to simulate how fluids move [5]. The tool, named the Artificial Intelligence driven air-Safety Tool (AISaT) [6], is being developed to learn from indoor airflow and provide recommendations for the placement of mitigation devices, such as HEPA filters. The tool is intended to identify the best position for the HEPA filter so it can be the most effective at removing aerosols from that space, with the overall goal of preventing transmission of respiratory infections. The research team are currently conducting a randomised control trial (RCT) with AISaT in the hospital setting, and usability testing sessions with the AISaT were carried out before using it in the RCT.

Whilst AI has the potential to enhance services and optimise resources within healthcare, considerations need to be made to improve the likelihood of its uptake and long-term use [7]. A key facilitator to the implementation of AI within hospital settings is to involve end-users in the development of the tool [8]. The involvement of end-users can be achieved through usability testing, which is a technique to capture the perspectives among users of a technological innovation as they interact with it [9]. This manuscript presents the findings from a qualitative usability testing study with hospital staff, which aimed to use their views of an early-stage version of the AISaT user interface, to enhance its usability and likelihood of integration into existing workflows.

## Methods

A qualitative observational study was conducted following Health Research Authority ethical approval [25/EM/0156]. The study design factored in the iterative sharing of emerging findings with tool engineers, allowing the engineers to consider the updates for a new version of AISaT.

### AISaT

The AISaT Mock V2.0.0 interface was used during the usability testing sessions (Figure 1). This version of AISaT is split into six steps or stages representing a consultation room in a hospital, these six stages have been visualised in Figure 2.

**Figure 1.**
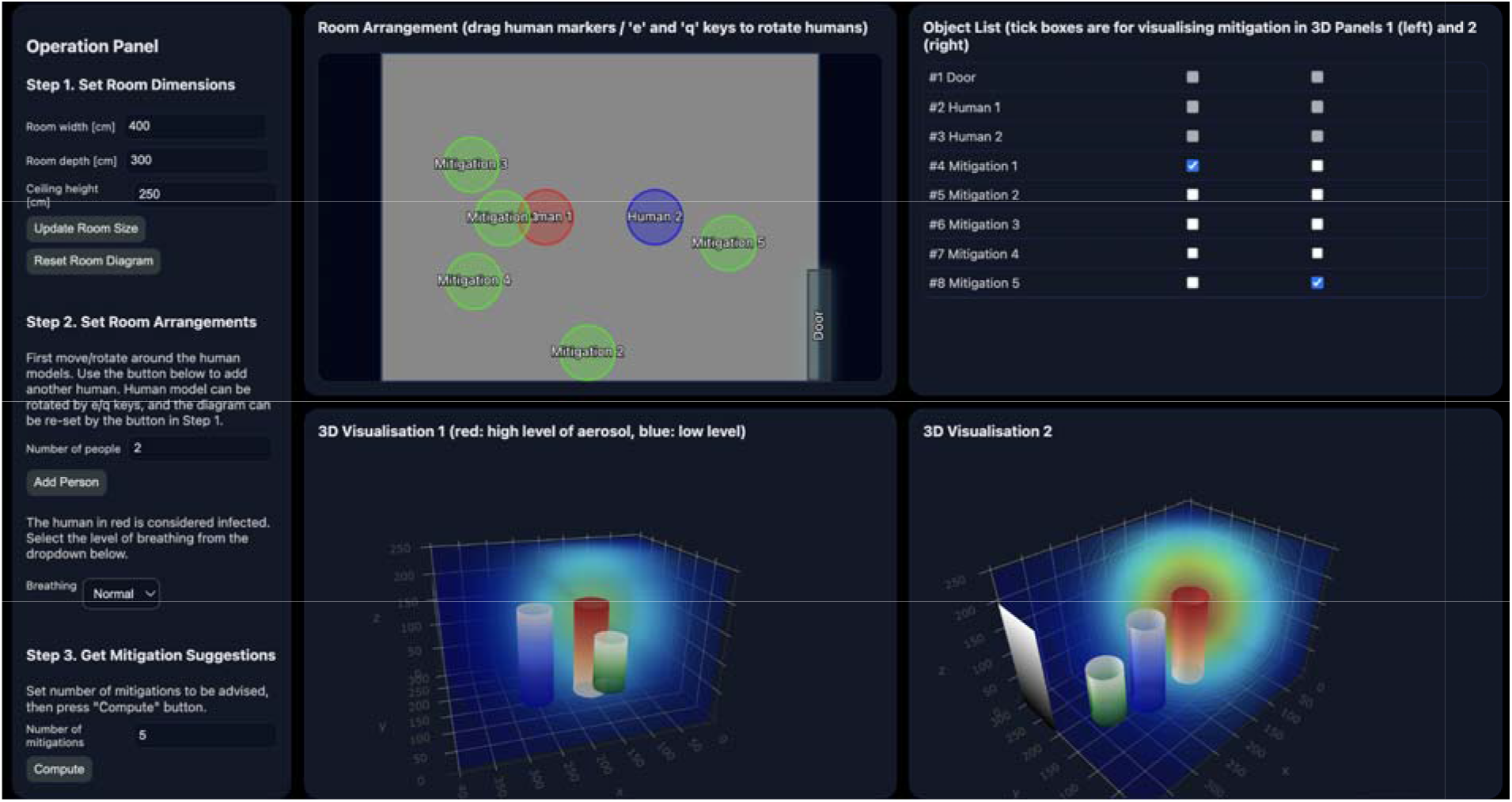
Interface of AISaT Mock V2.0.0.

**Figure 2.**
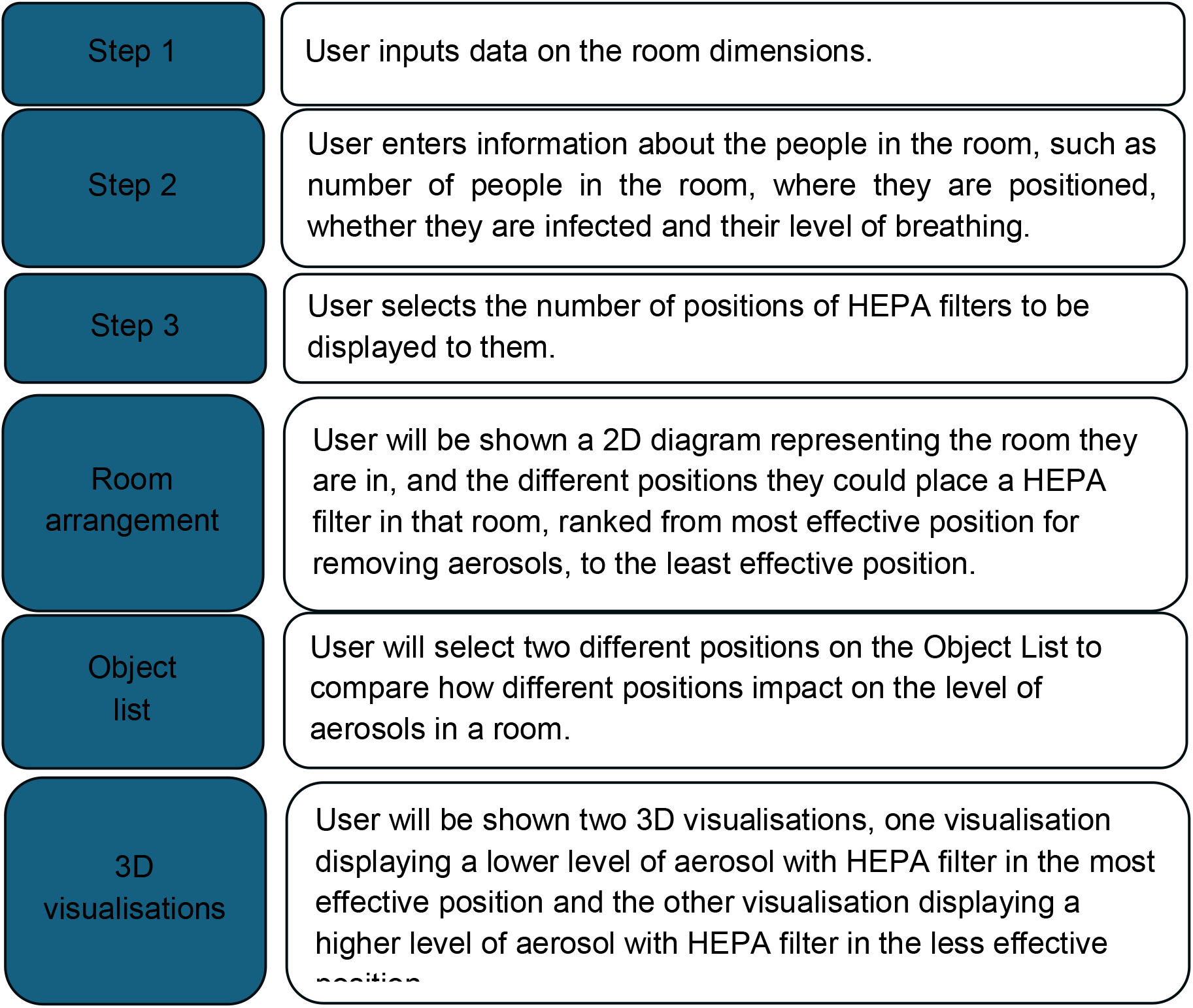
Stages of the AISaT Mock V2.0.0 interface.

The study was conducted with staff across two hospital sites in London. Convenience sampling was conducted at the sites, where a local stakeholder was asked to introduce the research team to colleagues with expertise in clinical activities, infection prevention and control (IPC), or estates management. These specialities were selected as they would most likely be responsible for using AISaT in a real-world scenario. Staff from these groups will likely be responsible for inputting details on the room dimension, on the participants in the room, or acting on the recommendations provided by AISaT in terms of positioning HEPA filters. An informed consent process was used with all participants, and everyone signed a consent form.

### Data collection

The usability testing was conducted with AISaT Mock Version 2.0.0 between November 2025 and May 2026. AISaT was loaded onto a Google Chrome page through a remote server at University College London (UCL), as only UCL staff could access the webpage. The researcher (S.E.C) loaded the webpage up on her own device and displayed the page via Microsoft Teams to the participant. The researcher demonstrated AISaT to the participant describing each of its components, rather than the participants using the tool directly themselves. Following the demonstration, the researcher asked the participants to ‘Think Aloud’ describing their thoughts on the demonstrated AISaT interface. The researcher also went back through each component and asked the participant prompting questions. During the usability testing, field notes were developed, an audio recording of the discussion was made and fully transcribed.

### Data analysis

The field notes were initially analysed using RREAL sheets [10] to visualise and chart the data. The RREAL sheet collated data on the different steps within AISaT, the overall interface, opinions on ownership, and how frequently it should be used. The RREAL sheet was then used to share updates and recommendations with the engineering team in real-time as data was being collected.

Framework analysis was then conducted with the transcripts [11], which were charted into the deductive themes listed below based on the adaptation of a usability testing framework developed by Richardson et al. (2017) [9].

- **Understand-ability:** whether the user can understand the instructions and purpose of each component in the tool.
- **Content:** whether the user thinks the tool is providing accurate and appropriate information.
- **Navigation:** whether the user would be able to move between different parts of the tool easily.
- **Visibility:** whether the user would be able to recognise key outputs from the tool.
- **Usability:** whether the tool could be used with minimal effort.
- **Workflow:** whether the tool could integrate easily into the user’s usual routine.

Additional deductive thematic areas were added to the framework to capture data related to ownership and how frequently it should be used. Inductive themes were also identified from the data.

## Results

### Sample characteristics

Ten participants across the two sites were interviewed, from professional roles: in clinical areas (n=6): estates (n=2); and IPC (n=2). Similar sample sizes have been used to assess the usability of tools in previous research [9].

### Findings from the usability testing

**Table 1.** Thematic areas identified from usability testing interviews.

| Themes* | Sub-themes |
| --- | --- |
| **Understand-ability**<br>*Whether the user can understand the instructions and purpose of each component in the tool.* | • Level of instruction throughout AIsaT<br>• Instruction around the mitigation devices<br>• Renaming components for better understand-ability |
| **Content**<br>*Whether the user thinks the tool is providing accurate and appropriate information.* | • Content about infection<br>• Content about the room<br>• Content about positioning devices<br>• Content about the object list |
| **Navigation**<br>*Whether the user would be able to move between different parts of the tool easily.* | • Page positioning<br>• Navigation of items in the room arrangement figure<br>• Navigation of 3D visualisations |
| **Visibility**<br>*Whether the user would be able able recognise key outputs from the tool.* | • Overall interface visibility<br>• Visibility of the room arrangement figure<br>• Visibility of the 3D visualisations |
| **Usability**<br>*Whether the tool could be used with minimal effort.* | • Overall usability<br>• Challenges interpreting 3D visualisations<br>• Challenges interpreting the object list |
| **Workflow**<br>*Whether the tool could integrate easily into the users usual routine.* | • Accessibility of the room measurement data<br>• Accessibility of data on the people in the room<br>• Mode of use<br>• Real-life positioning of devices |
| **Ownership**<br>*Who would likely be responsible for using the tool.* | • Ownership of finding out room measurements<br>• Overall ownership of using AIsaT and |
|  | positioning devices |
| Frequency of use<br>*How often the tool could be used.* | • Weighing up level of accuracy against feasibility |
| Inductively identified thematic areas<br>*Topics found in the data that were not part of the original framework* | • A more advanced version of AISaT<br>• Concerns with future mitigation measures incorporated into AISaT<br>• Saving outputs for audit trails |

\*Adapted from Richardson et al. (2017) [9]

### Understand-ability

#### Level of instruction throughout AISaT

<u>P</u>articipants (n=5) wanted further instruction throughout the process of using AISaT, to understand what the user is expected to do at each step in greater detail. Specific areas highlighted included ‘Step 2 – Set room arrangements’: users would need more guidance on how to move the human avatars within the room arrangement figure, and instructions that the items in the figure need to reflect the position of things in the room in real-life. Participants also needed further detail on the ‘Object list’, so users knew this was linked to the ‘3D visualisations’ component. Headers in the table could make it clearer the user is comparing two device positions. The addition of labels to the ‘3D visualisations’ with an explanatory paragraph about the difference in level of aerosols between the two visualisations would also be helpful.

> *“But then again, the wording isn’t so clear, what you’re asking us to do. The wording is saying they [humans in room arrangement figure] can be rotated, but really*…*what you want us to do is to rotate them to how they are in the room*.*” A02*
>
> *“I wonder if at the top of the two tick boxes, you could just have like left, right, [visualisation] rather than it being one long sentence?” B01*

#### Instruction around the mitigation devices

There was a lack of clarity on what the user was supposed to do with ‘Step 3 – Get mitigation suggestions’, further instruction was recommended, to tell the user they are being shown five different positions to choose from, rather than that the user needs to place a HEPA filter in all five of the positions. Guidance was also needed to explain that the positions would be ranked from most effective (position 1) to least effective (position 5).

> *“When you selected the mitigation suggestions, it wasn’t clear whether I was meant to be having five different air safety filters to be placed in those five places or those were just suggestions we need to place one*.*” A02*
>
> *“Is position one the best position to the next best down to position 5 being the least effective?*… *Is there some way of making sure that the user of this knows that?” A04*

#### Renaming components for better understand-ability

Participants suggested re-naming items within AISaT, veering away from using the terminology ‘human models’ and instead using phrases like ‘human avatar’ or ‘human 1 and human 2’ or ‘patient and healthcare professional’. The description of ‘mitigations’ throughout AISaT was also challenging to understand, participants suggested re-phrasing this to ‘position of mitigation device’ or ‘most effective position of mitigation device’, or even naming the specific mitigation device such as HEPA filter.

> *“I think even like sort of human one and human two could just be like patient and then HCP healthcare professional*.*” B01*
>
> *In real life, when it’s real, will it say mitigation one is a HEPA filter or mitigation 2 is a fan? Will it say that because, or will people have to just say it’s any mitigation thing in one of those positions?” B03*

### Content

#### Content about infection

The appropriateness of the content in relation to infection within AISaT was discussed. Some thought that AISaT should assume all patients were infected and all HCPs were non-infected as a model to reduce staff sickness and to prevent staff acquiring infections from patients and passing infections onto other patients. However, others raised concerns that staff should be considered infected too, as they could be a vehicle of transmission to patients. Additionally, automatically labelling all patients as infected may raise fears among HCPs. Moreover, the assumption that patients were all infected could lead to wasted resources e.g., more HEPA filters, or enforcing staff to wear PPIE unnecessarily.

> *“If you assume the patient always infected, you’d always be wearing an FP3 that unfortunately is incredibly uncomfortable, it does affect your performance… they may actually flag everything as a risk and everything can need to be filtered…with 10,000 machines everywhere that we cannot afford*.*” A06*

Participants thought it was important to have the option to select more than two people in a room if a carer joins a consultation, or in a procedure room or ward setting where there are multiple staff or patients. If more people were added, they would need to have the option to be made infectious or labelled as a patient or HCP. Some thought it would be important to define the specific infection such as respiratory infection, or even to include the type of respiratory infection as a variable that could impact the position of the HEPA filter e.g., Influenza vs COVID vs Tuberculosis.

> *“Depending on the viruses, for example, which patient you are having, like if it’s like influenza versus COVID versus measles versus TB…do you need like a different measure?” A06*

From an IPC perspective, level of breathing didn’t seem like a suitable variable among some participants, and activities such as coughing, speaking loudly or abnormal breathing seemed more appropriate. Whereas from a clinical angle, laboured breathing or coughing seemed appropriate. Two participants thought the option to choose the level of breathing should be removed from AISaT, and just assume all patients are breathing heavily for protection from the worst-case scenario.

> *“You’re going to put a filter in the room, you don’t know about the patients and the breathing because in any one day you could have infected, non-infected, low breathers, high breathers, so you’re not going to do this for every different patient that comes in*… *I’m going to assume they’re all infected and they’re all heavy breathers because that’s worst-case scenario, because otherwise, what’s the point of doing anything less?” A04*

#### Content about the room

Participants (n=7) thought AISaT was limited as it did not take into consideration existing variables within the room that affected air changes and ventilation. AISaT did not capture existing windows, or pressure differences due to corridors, furniture, screens, curtains, ventilation grills, or chill beams within the room. They recommended developing AISaT in line with the Health Technical Memorandum 03-01 (HTM 03-01) to first understand what air flow is like in a room. Also recommended was the integration of a device within AISaT that captures air changes in a room. Participants raised the need to find a way to factor into AISaT how people may move around the room. Another point that should be captured by AISaT is how the room is used. If it is a ward setting, the patients will be in the room a lot longer than a consultation room setting, and the surrounding areas in the room may have more infectious aerosols than in a consultation room. Likewise, it would be important to consider the type of procedure taking place in that room as ventilation requirements would be different depending on whether the procedure is invasive or non-invasive.

> *“I know it’s got a very basic room dimensions, but what if there’s other things in the room? Same clinics, we tend to have trolleys, couches, beds, etc*.*” B01*
>
> *“Without considering what’s happening to the air in that room in detail, you can’t really know where to put the device and your AI model needs to maybe consider that*.*” A03*
>
> *“You have to factor in other things like what is actually in the room currently, because that would, that might affect… airflow*.*” A07*

#### Content about positioning devices

Some participants liked having the flexibility to choose how many positions were offered for placing the HEPA filter, while others felt uncomfortable deciding how many options to display. Participants also raised that they would get frustrated if AISaT suggested unrealistic positions of placing devices such as in front of doors, far from plugs, or in front of a patient’s face. An alternative approach was suggested: where the user chooses where to place the device within the room layout based on their real-world knowledge of the space (for example, plug socket locations), and AISaT then shows how that placement would affect aerosol distribution. Other suggestions included highlighting different zones of the room with traffic light colours of the most to least effective positions. Also raised was the need for greater detail on where the device should be positioned in the room arrangement figure, such as the height or on certain furniture. Participants wanted more detail about the specific devices, such as the type of model of machine and how much it might cost. They also suggested including a percentage in reduction of aerosol next to each mitigation position throughout AISaT.

> *“I don’t want to see like ridiculous things like putting a HEPA filter right in between me and a patient’s face*.*” A01*
>
> *“I’m thinking if you may add amber colour to it. So we will have like a traffic light kind of warning*.*” A05*
>
> *“You need much more details… like a granularity of where do you really have to put the machine, what type of machine, how many air changes are you suggesting, what is the noise level?” A06*

#### Content about the object list

The inclusion of more variables within the object list was recommended, such as furniture within the room, and grouping together different variables such as furniture (chair, table and bed) and people (healthcare professional and patient).

> *“You could have like a little subheading, so split it into three boxes and then you can say devices and then you could put like fan, HEPA filter…instead of being one long list*.*” B01*

### Navigation

#### Page positioning

When it came to navigation across the whole page, one participant liked having AISaT labelled into steps, and another appreciated having the whole interface presented to them on one page. However, three participants had an opposing opinion that the whole interface presented on one page was overwhelming, they would prefer to scroll or click through each section sequentially.

> *“I prefer to be shown and told the steps one at a time not have all the steps and things that I’m meant to do available for me to see and look at because I think then it’s a bit overwhelming*.*” A02*

#### Navigation of items in the room arrangement figure

There was an appreciation that the room arrangement figure could be adjusted and tailored to reflect a user’s setting. However, some participants (n=3) found the navigation of human avatars in the room arrangement figure challenging as it relied on using keys on the computer keyboard, some suggested navigation using a computer mouse or cursor instead. They also raised there was not an option to position the door onto all four walls in the room arrangement figure which should be updated as the door location could affect air flow. One participant thought it would be useful to have a standard list of furniture items that users could click and drag into the room.

> *“I think the bit where it’s if it’s control Q and things to move the thing, then that’s a bit difficult*…*Rather than like using the mouse to wiggle them around a bit*.*” A01*

#### Navigation of 3D visualisations

Two participants thought the navigation within the 3D visualisations should be improved: adding instructions that the visualisations can be rotated by clicking and dragging on them; and enabling a function so the visualisations lock in the same pane, so when you rotate one visualisation, the other rotates too.

> *“Perhaps when you’re moving the first visualisation, the second visualisation is also locked in the same perspective, so then it’s…easier to compare*.*” B02*

### Visibility

#### Overall interface

Participants did not have concerns with the overall colour scheme of AISaT other than in relation to the room arrangement figure, but a couple of participants stated that the text throughout AISaT could be made larger.

> *“I think this works. I think it might just be a simple case of kind of making certain text bigger*.*” B02*

#### Visibility of the room arrangement figure

The participants valued having the visual of the room, and that the diagram was relatively simple, and that the different colours of the human avatars helped to distinguish between who was infected vs not infected. However, a couple of participants would have preferred if the figure was white instead of grey.

In relation to the room arrangement figure and the 3D visualisations, one participant suggested engineers may prefer to work in dimensions of meters rather than centimetres and suggested adding gridlines into the figure so users could work out the exact co-ordinates or distance from the wall of where to position the HEPA filter. Another suggestion included adding labels to items in the room such as patient, HCP, windows, or any equipment. Many participants highlighted issues relying on circles to represent humans or HEPA filters and suggested using more realistic graphics of a HEPA filter and other furniture. They also recommended using human avatars with faces that show the direction aerosols are being emitted. Another suggestion was to make it clear if humans are standing or sitting, and whether HEPA filters are positioned on the ground or on furniture.

> *“So, I would perhaps have it so an arrow or a smiley face or something so you can tell which way up your human is*.*” B03*
>
> *“…Having a grid on the drawing would be useful so that it’s got…something that people could measure and… work out where it would be…you’re just guessing where that is now, you know slight variation in position could make a big difference to where the filter needs to go, so the positioning needs to be as accurate as possible*.*” A04*

#### Visibility of the 3D visualisations

There was positive feedback on the 3D visualisations, that they were helpful to show the impact on aerosols of having HEPA filters positioned in different places. A participant highlighted that having colours in the room arrangement figure aligned to the 3D visualisation colours was helpful. Many participants said it was difficult to rely on colour to show differences in emissions, especially for people who are colour blind, and because the emission colours clash with the red and blue colours used for the human columns. Another participant highlighted when a HEPA filter (represented by a smaller column) was shown on the same pane as a human (represented by a larger column), the two columns merged in the visualisation and were hard to tell apart. In reality, one was positioned in front of the other, but this was only visible when the visualisation was rotated.

> *“That’s really quite powerful, isn’t it, to show the difference of placing the filter a slightly different position than the effect it has, or the importance of doing it?” A04*
>
> *“If you’re colourblind it wouldn’t be very helpful*.*” B03*

### Usability

#### Overall usability

Some of the participants thought AISaT seemed simple and quite user-friendly. But many highlighted that ongoing development of AISaT should ensure it is user-friendly and with limited content to read, with one participant suggesting a maximum time of 5 minutes to complete it.

> *“You want it to be as quick as possible because you’ve got lots of things to get ready. Before the list starts. So, you’d want this to be less than… 5 minutes, the whole thing from start to finish*.*” B03*

#### Challenges interpreting 3D visualisations

Users would likely have to spend some mental energy and effort to interpret the 3D visualisations. Specifically, due to the reliance on colour to interpret a difference in level of aerosols, the user would be expected to think through the colour wheel to work out whether there is a low or high number of aerosols. Participants thought AISaT should find another way to portray a difference in concentration of aerosols such as dots, circles, or actual numbers of aerosols. Concerns were also raised that if there were more than two people in the visualisation, it could get too busy and even more difficult to interpret. One participant suggested instead of relying on the quite mathematical looking visual/graph, impact of the specific position of HEPA filter on aerosols could be demonstrated with a picture of people in a room with loads of aerosols vs people in a room with minimal aerosols, highlighting within the picture that the HEPA filter is in a different place.

> *“The only real differences between those pictures is that the one on the right has a bit more yellow and green it well, not even yet. It’s got a bit more orange, but then I have to think right where would orange be on the red blue spectrum?” A01*
>
> *“[Demonstrate that] those two slides of between [the two positions]…getting it right gives you this protection. Getting it wrong means you’ve got no protection…so sort of try and visually show them the difference between what’s good and what’s bad…show them a picture*.*” A04*

#### Challenges interpreting the object list

Similar to some issues raised in the understand-ability section, one of the participants thought it would take a long time to interpret what the object list was demonstrating and that is was linked to the 3D visualisations.

> *“If I just read step three and I just looked at the object list, I would have to spend, I think, quite a long time playing around with it to start to understand what it was showing me*.*” A02*

### Workflow

#### Accessibility of the room measurement data

The participants would not know the dimensions of the rooms they work in but thought they could access this information by using a LiDAR application to measure the room, or consulting the ward manager, IPC, or estates team who may already know dimensions from floor plans. An estates professional however flagged that floor plans may not encompass the dimensions of every room in a Trust and another participant raised that not all hospitals have these floorplans. A couple of participants suggested that staff could physically measure the dimension of a room with measuring tape but flagged this would be challenging for non-square/rectangular rooms, also raised was that having to do this strenuous task could put users off from engaging with AISaT.

> *“I think generally speaking, and I’m probably speaking on behalf of everyone that kind of works in the wards, I don’t think…I would know the dimensions of the wards*.*” B01*
>
> *“Because what you really don’t want is the start of a thing someone says, the first question I don’t get the answer. I’m not doing it*.*” A01*
>
> *“I’d have to have some kind of measuring device…like that LiDAR app thing*.*” B03*

#### Accessibility of data on the people in the room

Many thought it would be easy to predict where people were likely to be positioned in a consultation room setting, based on positions of existing furniture, and therefore where people would be seated. However, they wouldn’t be able to predict whether a carer joins a patient, or where wheelchairs may be positioned in a room. Also raised was that in practice clinicians won’t always work in the same rooms, and therefore would not know the setup of the room until they were in it for the consultations. Two participants also discussed that it would be challenging to predict where people would be positioned in settings outside of the consultation room setting, so in Aerosol Generating Procedure rooms and on the wards, unless you relied on where beds were located as hotspots.

Most participants didn’t think they would know in advance whether a patient was infected unless already captured in their medical notes, and the user had time to read these before the appointment. One participant thought this data would be captured about patients on internal databases, and another suggested users could ask all patients in advance whether they were infected or had symptoms. Similarly, most would not know the level of breathing of patients until they saw the patient themselves, unless respiratory issues were documented in clinical notes. One participant also flagged that this component may make users think they need to measure the patients breathing, and if so, a standard measurement should be listed within AISaT.

> *“We know where people are going to sit, if it’s like a clinic room…well you sometimes get surprised…sometimes go out into the waiting room to collect your patient and you then realise they’ve got a family member with them. But generally, even if it’s two people I know where they’re sitting because I put the chairs out…you might find someone with a disability in a wheelchair and then you need to reconfigure your room*.*” A01*
>
> *“How do you know how infectious someone is before they come in? When I’m thinking of a clinic perspective, I guess if they were on the ward, you probably know how infective someone was given their symptoms*.*” B01*
>
> *“Yeah, I guess we’d probably need more clarification with what defines normal and what defines heavy. And I guess if they’re defining it based on the respiratory rate per minute, but then once again, it’s quite hard to make that call…before you see the patient and during the simulation*.*” B02*

#### Mode of use

AISaT could most likely be used on NHS computers according to the participants, however many flagged how slow NHS computers are. Another participant flagged that there may be barriers with NHS IT approving use of a third-party software. There were recommendations to host AISaT as a desktop application rather than a URL, so AISaT automatically opened or the application icon was made easy to find on the desktop. Some participants also suggested using AISaT on an iPad or iPhone so it could easily integrate with the LiDAR application and take photos of the room to calculate the dimensions.

> *“If it was quite basic, I think the NHS computers would be able to handle it. But I feel like…there’s so many things…that sometimes stop working or slowed down and whatnot*.*” B01*
>
> *“I guess from a look in a future application, I think it might be best where there’s an actual kind on the desktop, a desktop icon, so then they can just click on that and then it just pops up*.*” B02*
>
> *“Thinking about future developing of the app, you may want to consider maybe to integrate that into, for example, like an iPhone or an iPad. And then, you know, like you can take measurement very easily just with a laser*.*” A06*

#### Real-life positioning of devices

In terms of integrating the positioning of HEPA filters into a user’s workflow, participants raised concerns that some of the recommended positions may not be anywhere near plugs, and so their cables may create a trip hazard, or they may not be able to position the device in the location recommended by AISaT. Another concern was around the space available in rooms, some consultations rooms may only have two plugs available, and therefore not feasible to plug in the device. Similarly, AGP rooms are already crowded with equipment and people, and one participant thought it was unlikely you would be able to fit a HEPA filter into the room, let alone have numerous positions to choose from to position the HEPA filter.

> *“I’d still be thinking, even though it’s telling me this is the best place, I’d still be thinking, is that practical? Is…someone going to trip on it…it’s all very well it’s telling me that, but I can’t actually stretch the plug that long. So, it’s going to have to go here and that’s the best I can manage*.*” B03*
>
> *“You have to take into consideration the current layout in the room and whether putting that mitigation adds another risk… for example, falls and trips, if you have to have long cables for that mitigation to be in place*.*” A07*

### Ownership

#### Ownership of finding out room measurements

In terms of finding out the measurements of the rooms, there were dividing opinions on who could be responsible. Some clinical staff thought the estates team could support with completing this data as they may have it on file already, or they may have more experience in measuring rooms. One of the estates professionals thought this step could be completed by estates and IPC professionals, as they likely have this data already. One clinical participant thought AISaT could be configured so that all rooms in a hospital have been measured and pre-loaded into AISaT, perhaps by an estates professional, so the individual users who may be clinical staff, could then select their relevant room from a drop-down menu. They did reflect however that this would likely be a large time burden on the estates professional. A couple of clinical staff suggested the matron in charge or ward manager could find out this data and input the data for the relevant rooms they manage. Similarly, an estates professional felt like anyone could manage the measurement of a relevant room.

> *“I would hope that it would be somebody else, like the nurse in charge who’s got kind of oversight of the clinic setups and the and then knows the room dimensions and could do it all*.*” A01*
>
> *“Trust estates should have it…we’ve got square foot square area of what, you know each space*…*I’m sure IPC have a version of that too*.*” A03*
>
> *“So there’d have to be some way of measuring easily…do they send them a device to do that? Do they provide the app?*…*But then it’s another step, isn’t it?*…*So that’s why I think if the rooms were pre-measured already, say by estates…then that would take out that step for the clinical people and they’d be more likely to use it*.*” B03*

#### Overall ownership of using AISaT and positioning devices

When it came to overall ownership of AISaT and positioning of the devices, some clinical participants (n=2) suggested this could be carried out by the ward manager or matron and could be checked occasionally by clinicians using the room. They thought the clinicians themselves may not have time to use it whilst getting ready for the clinic and conducting consultations.

> *“Maybe the nurse in charge comes round and does a like a round of all the rooms and says…all your mitigations are in the best possible places*… *And then I go into my room and be like, well, it’s sitting there on my desk. It’s a bit annoying, so I’ll ask the tool or the nurse in charge. What’s the second best place? And then see if that’s something tolerable*.*” A01*

Some participants suggested that an estates professional could be responsible for AISaT and the placement and maintenance of devices, but if so, the language within AISaT may need updating, specifically around level of breathing which may be more clinical. A slightly opposing viewpoint was shared by an estates participant, who thought if an estates user were to use AISaT for all rooms, it would be a lot of work for one person, versus little work for many people to complete AISaT based on the individual room they work in. The participant also raised that an estates professional may have a generalised understanding of the room dimensions and how the room may roughly be used so long as furniture doesn’t move. But ultimately, it’s up to the clinician in the room how they setup the furniture, and they would most likely know more about their patients circumstances to input data on them into AISaT. Both opinions tie to a point shared by an interview participant, who said that the ownership would impact how frequently AISaT is used, and the accuracy of AISaT: The estates professional could take on the responsibility of inputting data based on room dimensions, and a generalised understanding of where people would most likely be positioned based on furniture. However, they may not be able to provide as accurate information into AISaT as the clinician who could use AISaT between appointments based on number of people in the room or whether they are infected. Similarly, but unrealistically, the clinician could even use AISaT during the consultation based on where people are moving within the room and change the positioning of the HEPA filter.

> *“So, you could go from 1 extreme and…get someone from estates or the nursing team who usually arrive and organise some things before the clinic to do this and leave it for the health clinic. Or the other extreme is…every single patient who comes in, and even within the consultation, the doctor is reassessing, you know, once or twice during the consultation and moving it. I think that extreme is highly unrealistic and impractical…would have to consider sacrificing I think accuracy or methodological ideals for practical*.*” A02*

A few participants suggested AISaT should be used in collaboration with a combination of the different staff roles: estates, IPC, IT, and clinical staff. With a couple of participants emphasising that before AISaT can roll out and HEPA filters positioned in certain rooms within a trust, it will be vital to get sign off from the Trust, IPC, the finance team, and the fire and safety team. To facilitate this, three participants suggested that there is one named lead stakeholder in charge of engaging and progressing the implementation of AISaT.

> *“I think the IPC would probably come to someone in estates and we would accompany them to that space, and if there’s a measurement issue, we could figure that out*… *But they [IPC] would decide where is the best place to put any device that might impact have a clinical impact on patients or staff*.*” A03*
>
> *“Say it’s showing that we need HEPA filters… 100% estates, finance, infection control would all want to be involved in that. And it wouldn’t be…just down to the ward manager to say, yeah, we’re going to buy four HEPA filters for this ward*.*” B03*

### Frequency of use

Most of the participants (n=8) thought it would be most feasible to use AISaT once, or for each morning or afternoon block of appointments, or only if furniture in the room changes, as opposed to in between appointments or during appointments. Alternatively, users could practise using the AISaT with different numbers of people in the room and learn the recommended HEPA filter positions, so they can place the filter based on memory, based on the number of people present on the day. Similarly, the user could memorise the best position based on whether the patient is infected or breathing heavily.

> *“You’re either going to do it at the beginning of the session and have it for everyone on that session and assume everyone…needs the same mitigation… it’s then it’s going to be difficult to do it in between patients if it was a heavy breathing patient*.*” B03*

### Inductive thematic areas

#### AISaT to be more advanced

A few participants (n=4) thought AISaT would need to be made more advanced to factor in more variables about the items within a room. These variables could include how many people are present (especially considering a ward setting) and how they move around within a room, the different types of infection they may have, the existing furniture, and ventilation and air change conditions. One of these participants raised that including AI in the name of AISaT, may lead to overly high expectations of what it can deliver, and the current version may not match those expectations.

> *“I think just already the name AI-based, you kind of set a different expectation, that isn’t, I mean, yes, it is AI, but it actually doesn’t really…do much of the work for you…it is a software to tell you eventually which position, but you still have to input quite a lot on that*.*” A06*

#### Concerns with mitigation measures incorporated into AISaT

One participant raised their concern that a HEPA filter would not be powerful enough for use in operating rooms which require a high number of air changes per hour. Another participant flagged their concerns with adding other mitigation measures beyond just recommending HEPA filter positions. They were concerned with devices like ventilated patient isolation tents, which may be successful in terms of air filtration, but are disempowering and uncomfortable to patients as it stops them being able to move around easily.

#### Saving outputs for audit trails

One participant thought it would be helpful if AISaT has a feature that allows you to save the information you have input and the recommendations on positions AISaT provided, as an audit trail.

## Conclusion

This study demonstrated that usability testing using qualitative research methods has been a helpful approach to capture key insights from professionals who may use AISaT in the real-world setting and inform improvements. Some improvements included renaming components to improve understandability, improving the navigation within the AISaT, and making updates to the visibility and usability of the room arrangement figure and 3D visualisations. The research team will need to critically reflect on the content within AISaT, largely around capturing more variables related to the room, and determining who is infected, and the level of breathing.

## Data Availability

All data produced in the present study are available upon reasonable request to the authors

